# Postal sputum samples for clinical and research applications in chronic lung infections caused by Pseudomonas aeruginosa

**DOI:** 10.64898/2026.06.27.26356742

**Authors:** Louise Jackson, Rosemary Maher, David Green, Warwick Dunn, Catherine Winder, Dharun Senthil Kumar, Wanchang Lin, Ed Emmott, Rebekah Penrice-Randal, Victoria Shaw, Susan Holden, Phil Mitchelmore, Ilona Littler, Kamlesh Mohan, Dilip Nazareth, Dennis Wat, Dan Wootton, Jo Fothergill, Freddy Frost

**Author notes:** **Correspondence** Dr Freddy Frost, Institute of Lifecourse & Medical Sciences, University of Liverpool, William Henry Duncan Building, Liverpool, UK.

## Abstract

**Introduction:** Postal sputum sampling represents a potential strategy for patient-led, efficient, and regular sampling, yet little is known about the validity of posting samples for clinical and research purposes in bronchiectasis. This study aimed to validate postal sputum sampling for clinical and research applications in chronic *P. aeruginosa* infection.

**Methods:** Sputum was collected from 12 participants with bronchiectasis and known *P. aeruginosa* infection. Each sputum sample was divided into four aliquots: two were sent immediately for analysis with or without ‘DNA-Shield’ (shield-fresh and non-shield-Fresh), while two were transported through the UK postal service, with or without ‘DNA-Shield’ (shield-posted and non-shield-posted). All aliquots were sent at ambient temperature and subsequently processed for bacterial enumeration through selective culture, detailed antimicrobial susceptibility testing, quantitative PCR (qPCR), 16S microbiome sequencing, metabolomics, and proteomics.

**Results:** During postage, there was a median of four days (range, 2-7) between sample collection and processing. 7/12 patients were positive for *P. aeruginosa* by culture of fresh samples, with 100% agreement in posted samples. Postage did not affect cultured (p=0.81) or amplified load of *P. aeruginosa* (p=0.94), and no differences were observed in AST profiles across 140 isolates for *P. aeruginosa* cultured from fresh or posted samples. Metabolomics and proteomics revealed that variation between individuals was significantly greater than between fresh and posted samples, and no significant differences in microbial taxa were observed between samples. No differences were associated with the addition of DNA Shield by qPCR (p=0.19), however, freeze-thaw from -80°C increased amplified load (p=<0.01).

**Conclusions:** We found little evidence of an effect of postage on sputum positivity, recoverable load, AST profile, microbiome, proteome or metabolome in sputum samples. These data suggest postal sputum samples may be a valuable tool for clinical and research applications.

## Introduction

The identification of key pathogens, such as *Pseudomonas aeruginosa,* in people with chronic lung infection relies heavily on sputum sampling. Clinical microbiology protocols often mandate fresh or same same-day sputum sampling, with refrigeration recommended for only short periods (1, 2). This results in limited opportunities for sample collection and processing, often relying on face-to-face clinical interactions. Postal sputum samples offer a potential solution, offering an efficient, near patient, and regular sampling opportunity that could be utilised in both a clinical and research setting.

In the UK, postal sputum sampling is being used by some services; however, this is yet to be widely implemented into clinical practice as the effects of postage on sample validity are not fully understood. In prior work, the stability of next-day postal sputum from CF patients was demonstrated for the recovery of six common species, including *Pseudomonas aeruginosa* (3). Similarly and more recently, pathogen abundance and diversity within sputum from CF patients was found to not be impacted by postage (4). In bronchiectasis, a 2010 study into the effect of processing time revealed that cultural load of *P. aeruginosa* was not significantly affected 48 hours post expectoration if sputum was stored at 4°C. However, a small but significant difference was observed in sputum left at room temperature (25°C) (5). Beyond this, little is known about validity of postal sputum for the detection of *P. aeruginosa* in bronchiectasis patients and people with CF receiving modulator therapy. Particular evidence gaps exist for understanding the impact of postage on novel multi-omic technologies. As the major opportunistic pathogen in people with bronchiectasis and CF, *P. aeruginosa,* is fundamentally important in chronic lung infection, both in a clinical and research setting. Therefore, this study aimed to determine the validity of postal sputum for clinically relevant outcomes and potential research applications in bronchiectasis, with a focus on *P. aeruginosa*.

## Materials and Methods

### Study Design

The Postal Sputum Testing for Early Detection (POSTED) study included adults with bronchiectasis attending Liverpool Heart & Chest Hospital NHS Foundation Trust, Liverpool, UK between December 20023 and February 2024. The objective of this study was to determine if postage has an impact on sputum, for clinical and research purposes. Samples were collected prospectively and then split and then either processed fresh or after postage.

### Study Participants

Participants were eligible if they had 2 or more sputum samples culture positive for *P. aeruginosa* in the last 24 months. Participants were eligible regardless of aetiology, but those with CF were included if they had been established on CFTR modulator therapy for >3 months. POSTED was an approved sub-study of the Liverpool University Biobank (Health Research Authority (18/NW/0771)).

### Sample Collection

Spontaneously expectorated sputum was split in four equal aliquots by clinical laboratory staff: Aliquot 1:raw sputum for same-day fresh analysis; Aliquot 2: raw sputum for postage; Aliquot 3: sputum with 2 mL of DNA/RNA shield (Zymo Research, Irvine, USA) for same-day fresh analysis; Aliquot 4: sputum with 2 mL DNA/RNA shield for postage. Same-day fresh samples were processed on the day of collection. ‘Posted’ samples were sent through the UK postal service adherent to regulations for shipping biological specimens (Royal Mail Group Limited, UK). All samples were transported at ambient temperature.

### Sample Processing

On arrival to the laboratory, sputum aliquots were weighed and homogenised with an equal volume of Sputasol (Oxoid, Thermo Fisher Scientific, UK). Samples were incubated for one hour at 120 rpm on an orbital shaker, before immediate culture and DNA extraction. Only non-shield sputum samples were processed for culture, with all four sample types undergoing DNA extraction and downstream processing (Figure 1). Non-shield sputum concentrations were adjusted with 1x PBS to account for the addition of DNA/RNA shield to the shield sputum samples.

**Figure 1.**
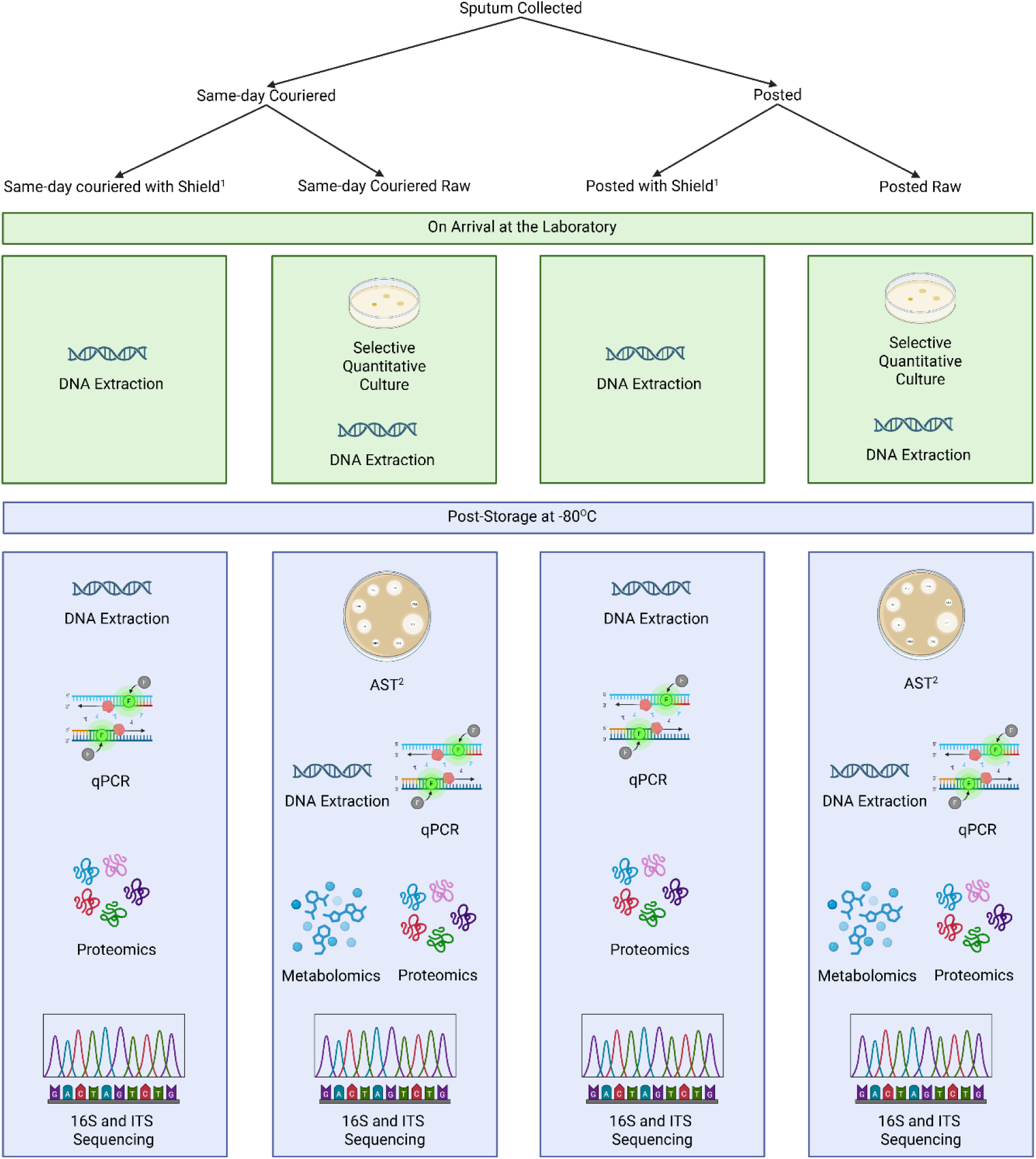
Flowchart of methods utilised in study. Same-day couriered samples were processed on the same day as collection. Posted samples were sent through the UK postal service (Royal Mail Group Limited, UK). ^1^DNA/RNA shield (Zymo Research, Irvine, USA) ^2^Antimicrobial susceptibility testing (AST) conducted on 10 isolates from each positive sample (140 isolates in total), against 12 antibiotics: Ciprofloxacin, Pip-Taz, Meropenem, Amikacin, Tobramycin, Ceftazidime, Aztreonam, Cefiderocol, Ceftazidime-Avibactam, Meropenem-Varborbactam, Gentamicin, Ceftolozane-Tazobactam.

**Table 1.** Baseline clinical characteristics.

| Demographic | N (%) or median [range] |
| --- | --- |
| **Participants** | 12 |
| **Sex** |  |
| Female | 5 (41.7) |
| Male | 7 (58.3) |
| **Age** | 47 [28-76] |
| **Exacerbations in last 12 months** | 4 [2-6] |
| **Hospital Admissions in last 12 months** | 2 [0-5] |
| **BMI** | 24 [19-29] |
| **Predicted FEV1** | 59% [27-77%] |
| **BSI** | 9 [7-11] |

BMI – body mass index, FEV1 - Forced Expiratory Volume in 1 second, BSI – bronchiectasis severity index.

### Bacterial Culture

A dilution series consisting of 200 µL of homogenised sputum added into 1800 µL of 1x phosphate buffered saline (PBS) was conducted, to produce a 1:10 dilution series up to and including 10^-8^. Dilutions were plated using the Miles and Misra method of enumeration (6), with 20 µL of each dilution spotted onto Pseudomonal-selective agar (Pseudomonas agar base with C-N Supplement, Oxoid, ThermoFisher Scientific, UK), in triplicate. Plates were incubated overnight at 37°C. A dilution with a countable number of colonies (2–200) was selected and average colony forming units were deduced. Three colonies were selected at random for confirmation of *P. aeruginosa* by matrix-assisted laser desorption/ionisation time of flight spectrometry (MALDI-TOF, Bruker, Germany).

### Antimicrobial Susceptibility Testing (AST)

Ten colonies per positive plate were stored in LB glycerol stocks at -80 for antimicrobial susceptibility testing (AST). A panel of 12 antibiotics were selected for AST (MAST, Bootle, UK): Ciprofloxacin (CIP5), Pip-Taz (PTZ36), Meropenem (MEM10), Amikacin (AK30), Tobramycin (TN10), Meropenem-Varborbactam (MEV30), Aztreonam (ATM30), Cefiderocol (FDC30), Ceftazidime-Avibactam (CZA14), Ceftazidime (CAZ10), Gentamicin (GM10), Ceftolozane-Tazobactam (C/T).

Disc diffusion assays were performed on bacterial lawns, produced using a 0.5 McFarland suspension of each isolate in 1x PBS on Mueller-Hinton agar. Zones of clearance were measured and compared to published guidelines of clinical breakpoints (7).

### Quantitative Polymerase Chain Reaction (qPCR)

Extraction of DNA was performed using the ZymoBIOMICS DNA Miniprep Kit (Zymo Research, Irvine, USA), as per manufacturer’s protocol for sputum. Primer sequences targeting the O-acetylase gene of *P. aeruginosa* (PA431CF 5‘-CTGGGTCGAAAGGTGGTTGTTATC-3’; PA431CR 5’-GCGGCTGGTGCGGCTGAGTC-’3) were lifted from Choi *et al*., 2013. The GoTaq qPCR master mix (Promega, Wisconsin, USA) was utilised and amplification was performed under the following conditions: 95°C for two minutes, followed by 40 cycles of 95°C for 15 seconds and 63°C for one minute, using the AriaMX Real-Time PCR System (Agilent, Santa Clara, USA).

### 16S Microbiome Sequencing

Full details of the 16S microbiome analysis are provided in the supplementary material. All sputum samples were extracted for V4 16S Illumina library prep. Libraries were sequenced on the Illumina MiSeq platform (Illumina, San Diego, USA), using V2 chemistry, generating 2 x 250 bp paired-end reads. Alpha and beta-diversity evaluations were conducted in Rstudio (v2025.05, Posit Software PBC) using the phyloseq, microbiome and vegan packages.

### Metabolomics

Full details of metabolomic analysis are provided within the supplementary material. All sputum samples were extracted applying a monophasic extraction/protein precipitation method and were subsequently analysed applying untargeted metabolomics assays using gas chromatography-mass spectrometry (GC-MS) and ultra-high performance liquid chromatography-mass spectrometry (UHPLC-MS) Full details are provided within the supplementary information.

### Proteomics

Full details of the proteomic analysis are provided within the supplementary material. After thawing, sputum samples were denatured with SDS, reduced with dithiothreitol and alkylated with iodoacetamide. Proteins were digested overnight with trypsin at 37 °C. Digestion was stopped by addition of trifluoroacetic acid and the samples were centrifuged to remove particulates. Tryptic peptides within samples were analysed by liquid chromatography mass spectrometry using an Evosep One (Evosep Biosystems, Denmark) coupled online to a hybrid trapped ion mobility spectrometry - quadrupole time of flight mass spectrometer (timsTOF HT, Bruker Daltonics, Bremen, Germany) with a modified nano-electrospray ion source (CaptiveSpray, Bruker Daltonics).

### Statistical Analysis

Where not otherwise described, statistical analysis was performed using GraphPad Prism 10 (GraphPad Software) for Windows, with heat maps and sensitivity/specificity data deduced using RStudio for Mac (version 2023.06.1+524) with the ‘pheatmap’ package (version 1.0.13, ). Statistical comparison between posted and fresh was determined using a Wilcoxon signed-rank test, with multiple Man-Whitney analysis to determine differences in AST profile.

## Results

### Patient Demographics

Samples were obtained from 12 participants. Patient demographics are outlined in Tabel 1). Forty-eight aliquots were successfully collected from all 12 patients.

### Postal Time

Postal samples took a median of four days (range, 2-7) to arrive at the laboratory. All samples (fresh and postal) were processed a median of 0 days (range, 0-1) post-arrival at the laboratory.

### Cultured Load

Of the 12 paired non-DNA Shield samples, 7/12 were positive for *P. aeruginosa* by culture, with 100% agreement in positivity/negativity between fresh and posted samples. In positive *P. aeruginosa* samples, no difference in cultured load was observed between fresh and postal samples (median (IQR)

6.0 Log_10_ CFU/mL (4.1-7.4) vs. 6.6 Log_10_ CFU/mL (6.2-7.2); p=0.81).

### AST Profile

No significant differences were observed in antimicrobial susceptibility profiles of 140 isolates of *P. aeruginosa* against 12 antibiotics, cultured from fresh or posted samples. For example, zones of inhibition were similar for all antibiotics tested, see Figure 3B. Similarly, when the EUCAST determined resistance was compared for all 140 isolates, almost identical patterns were observed (Figure 2). Whilst there are currently no published breakpoints for gentamicin against *P. aeruginosa*, the mean zone of inhibition did not significantly differ between fresh and postal samples (mean (SD) 10.4 mm (10) vs. 12.1 (10); p-value=0.97). Antibiotic sensitivity in isolates from posted samples had positive predictive value of 91.8% and negative predictive value of 88.2% for sensitivity in fresh samples. A Kendall’s tau was performed to determine correlation between postal and fresh sputum, and a strong associated was observed between zone of inhibition of isolates from postal and fresh sputum (R=0.74; p =<0.005).

**Figure 2.**
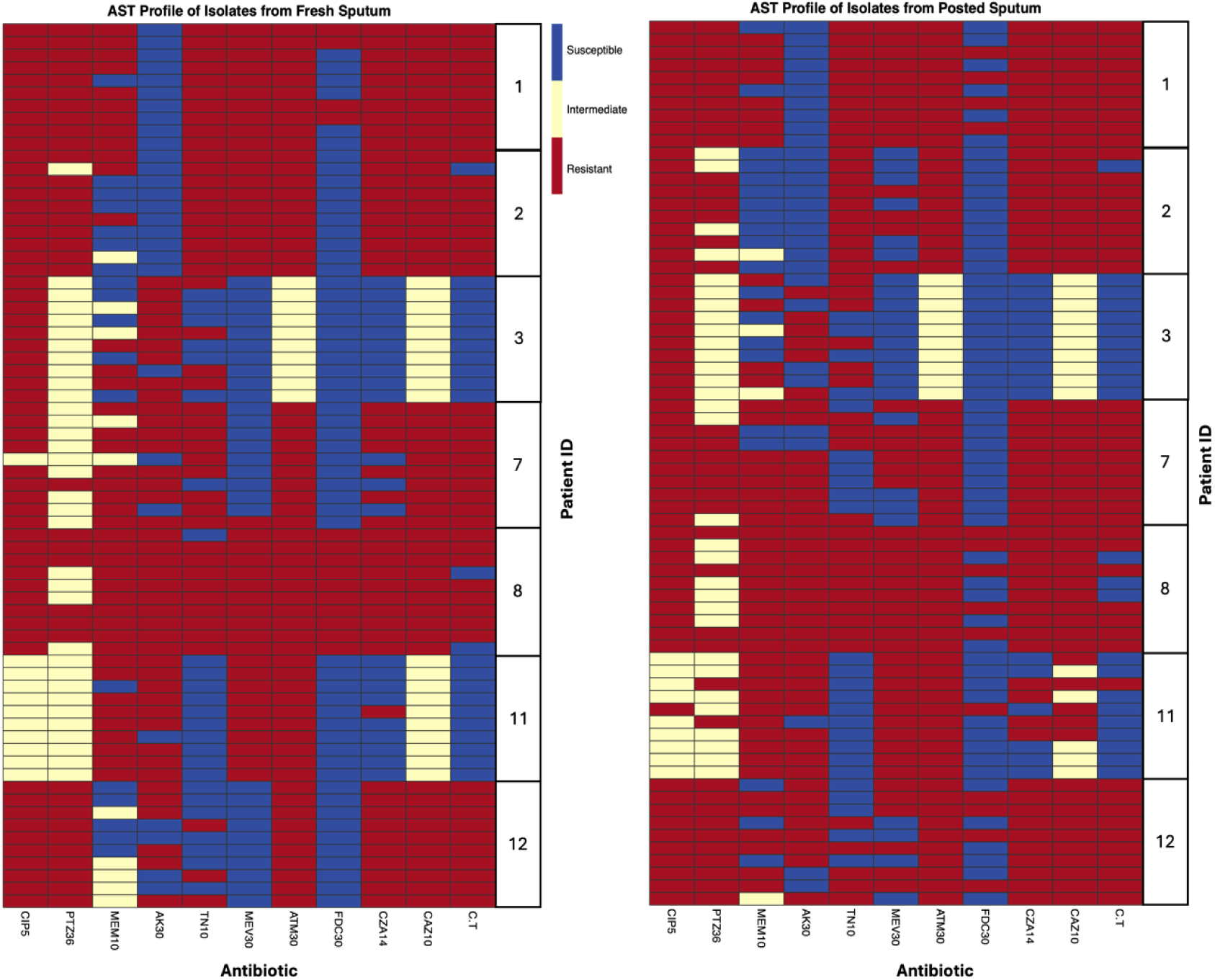
Heat map data for antimicrobial susceptibility profiles of *P. aeruginosa* isolates against 11 antibiotics^1^, from fresh and posted sputum of bronchiectasis patients. ^1^CIP5 - Ciprofloxacin, PTZ36 - Pip-Taz, MEM10 - Meropenem, AK30 - Amikacin, TN10 - Tobramycin, MEV30 - Meropenem-Varborbactam, ATM30 - Aztreonam, FDC30 - Cefiderocol, CZA14 - Ceftazidime-Avibactam, CAZ10 - Ceftazidime, C/T - Ceftolozane-Tazobactam.

**Figure 3.**
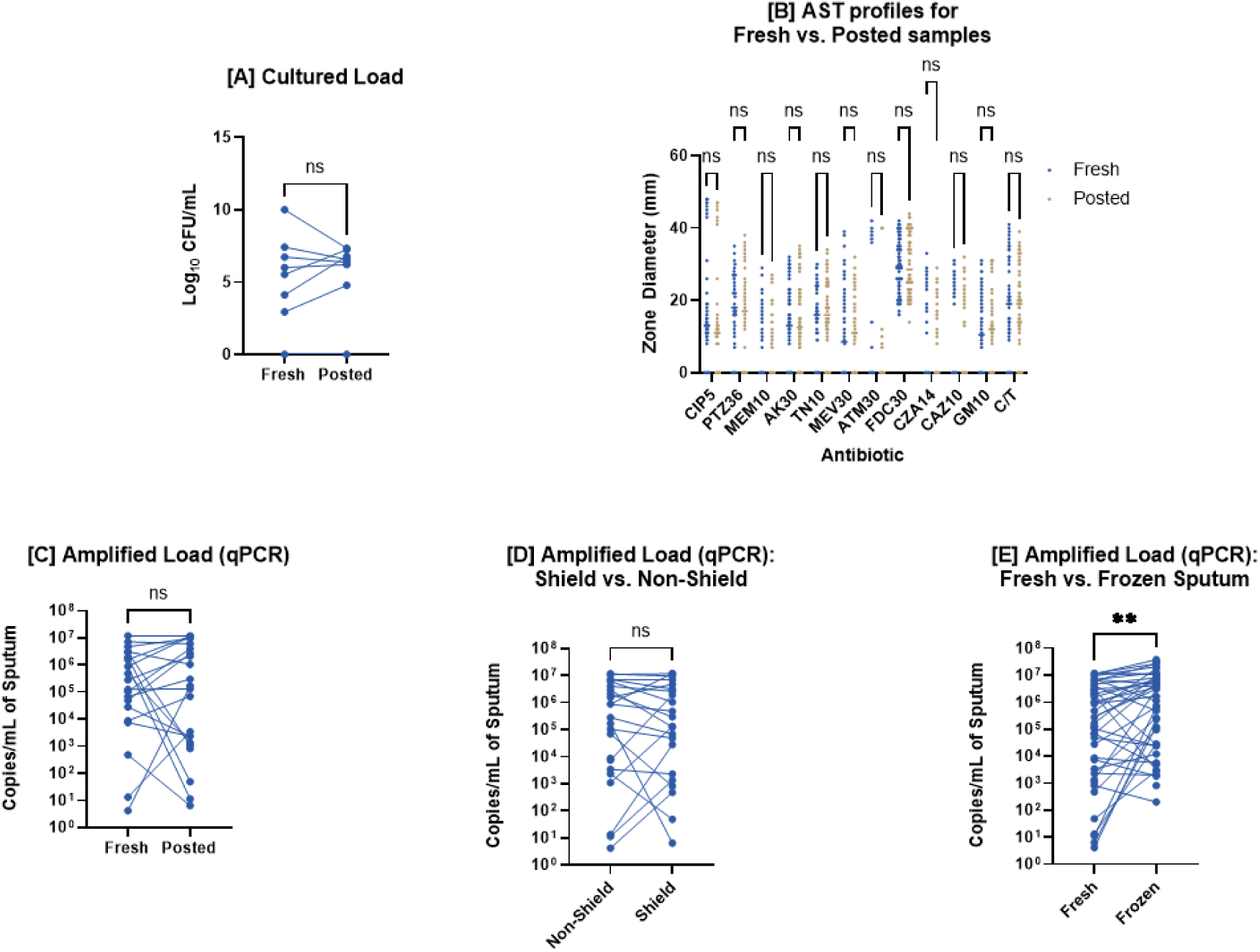
[A] Cultured load of *P. aeruginosa* from fresh and posted sputum samples^1^. [B] AST profiles for isolates from fresh and posted sputum samples, against a panel of 12 antibiotics^2^. [C] Amplified load of *P. aeruginosa* in fresh and posted sputum samples. [D] Amplified load of *P. aeruginosa* in sputum with the addition of shield and without shield (non-shield). [E] Amplified load of *P. aeruginosa* in sputum extracted fresh and post freeze-thaw at -80°C^3^. ^1^ Sputum samples were collected from patients with bronchiectasis ^2^CIP5 - Ciprofloxacin, PTZ36 - Pip-Taz, MEM10 - Meropenem, AK30 - Amikacin, TN10 - Tobramycin, MEV30 - Meropenem-Varborbactam, ATM30 - Aztreonam, FDC30 - Cefiderocol, CZA14 - Ceftazidime-Avibactam, CAZ10 - Ceftazidime, GM10 - Gentamicin, C/T - Ceftolozane-Tazobactam ^3^determined by qPCR (8)

### qPCR P. aeruginosa load

There were no significant differences in amplified load of *P. aeruginosa* (p=0.70) between fresh (median 8.3 Log_10_ copies/mL of sputum; IQR, 6.9-8.8) and posted samples (7.5 Log_10_ copies/mL of sputum; IQR, 5.4-9.0). However, post freeze-thaw DNA extraction of sputum from -80°C storage significantly improved amplified load (p=<0.01). No significant differences in amplified load of *P. aeruginosa* were detected between samples with and without the addition of DNA Shield by qPCR (p=0.19; Figure 3).

### 16S Microbiome Shequencing

Overall, after sequencing >10million reads in 48 samples, we found no change in the number of taxa that were identified between fresh and posted samples (Figure 4 [A]). Similarly, no difference in either Shannon or Fisher diversity metrics were observed (Figure 4 [B] & [C]). Overall, in terms of community composition we observed more variation between individuals than between fresh/posted samples. For example, samples from the same individual were generally closely related, regardless of posted/fresh status, and there was no clustering by posted/fresh status (Figure 4 [B] & [C]). Similarly, no taxa were differentially abundant between fresh and posted samples (Supp. Figure 1). PERMANOVA analyses suggested Patient ID explained 72% of all community composition variation compared to 0.9% explained by fresh/posted.

**Figure 4.**
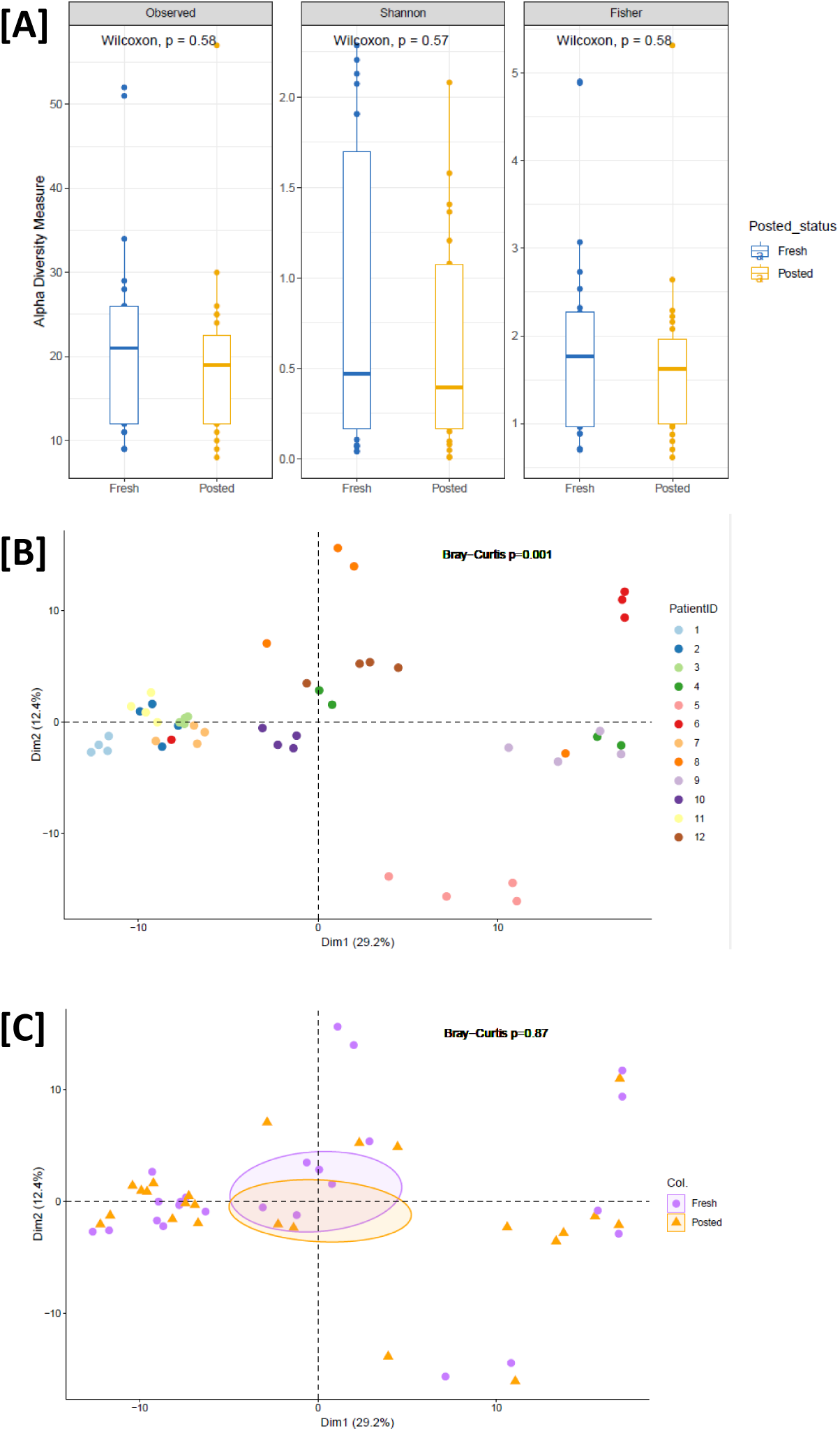
Alpha diversity metrics in posted and fresh samples. [A] A principal coordinate analysis (PCoA) of 16S rRNA gene microbiome data. [B] Each point represents a single sample, coloured by Patient ID showing close clustering of samples by participant confirmed by PERMANOVA (Bray-Curtis p=0.001). [C] A PCoA of 16S rRNA gene microbiome data, each point represents the same samples from B, this time coloured by posted/fresh group with overlapping 95% confidence interval ellipses. PERMANOVA confirming not significant differences between groups (Bray-Curtis p=0.87).

As expected, most patients’ microbiota were dominated by *Pseudomonas*, which accounted for 54.3% of all reads. We did observe significant differences in the abundance of *Pseudomonas* in those patients who were culture positive vs culture negative on the day for *P. aeruginosa* (median relative abundance 96.3% vs 3.4%; p=0.004).

In one patient (POST004), no *Pseudomonas* reads were observed in two posted samples despite >90% of reads in the fresh samples being attributed to *Pseudomonas*. Amplified load for *P. aeruginosa* confirmed no reads in the posted samples when analysed fresh. However, repeat extraction post-freeze-thaw cycle did confirm a sequenced load of *P. aeruginosa* in the posted sample equivalent to that in the fresh samples suggesting an extraction failure rather than impact of postage (Supp. Figure 2).

### Metabolomics

Untargeted metabolomics analysis of all 48 sputum samples was performed. A large dry mass was observed for lyophilised samples containing DNA shield. The mass chromatograms observed for GC-MS and UHPLC-MS analysis of samples containing DNA shield contained only a few chromatographic peaks and were significantly different to mass chromatograms for samples not containing DNA shield which contained a larger number of chromatographic peaks. Data acquired using GC-MS and UHPLC-MS for the 24 samples not containing DNA shield were processed further. No metabolites were statistically significantly when comparing non-shield fresh and non-shield posted samples (p < 0.05 after multiple testing correction with Benjamini-Hochberg method). Principal Component Analysis and Hierarchical Clustering Analysis demonstrated that technical differences between fresh and posted samples for the same participant were much lower than biological differences observed between different participants. Figure 5 visualises hierarchical cluster analysis (HCA) analysis for GC-MS and UHPLC-MS positive ion mode.

**Figure 5.**
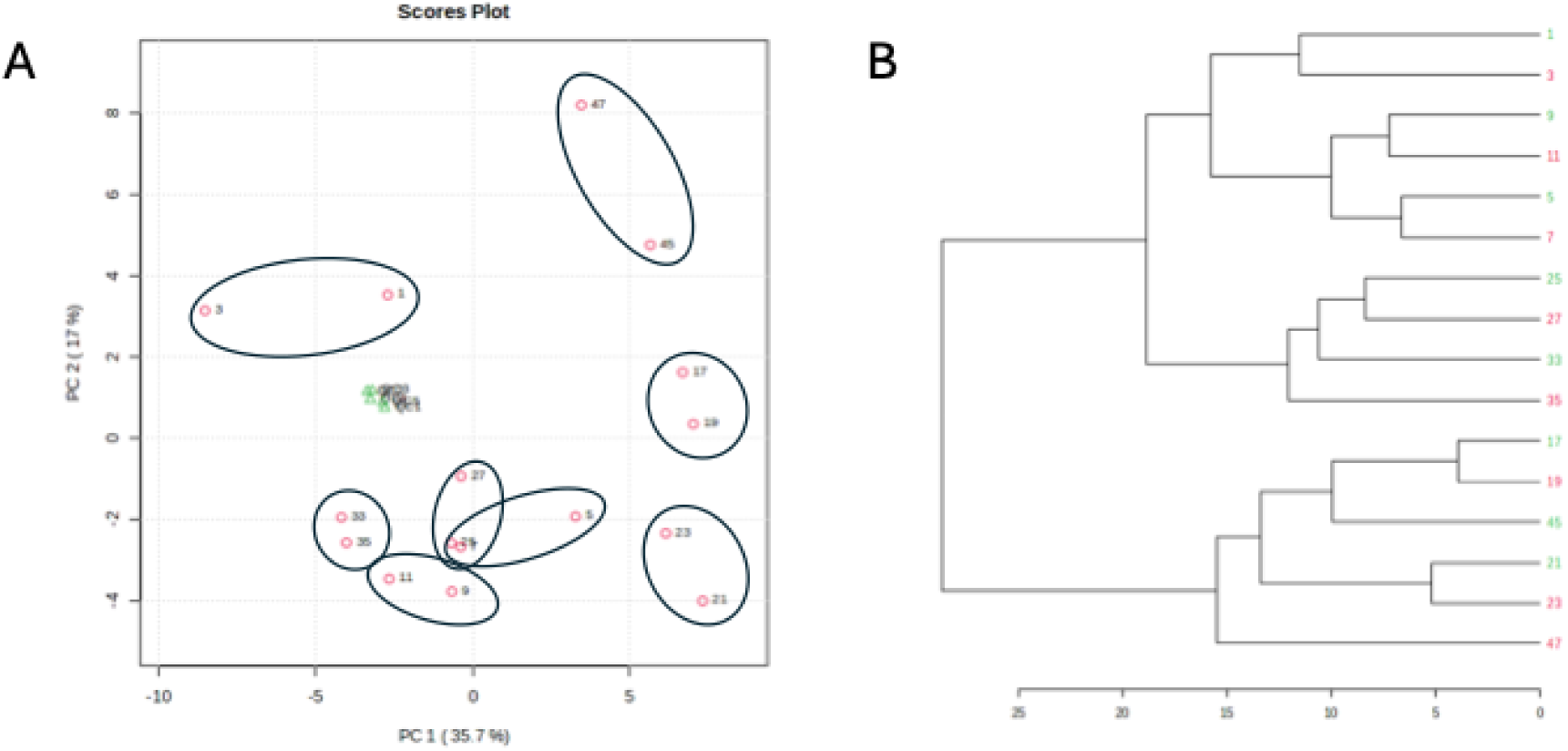
Untargeted GC-MS metabolomic profiles are stable between posted and fresh samples from the same patient. [A] Principal component analysis with each red symbol representing a sample, numbered by sample ID. Paired fresh and poster samples from the same patient are identified by ellipses. Green symbols represent the quality control (QC) samples. [B] Hierarchical Clustering Analysis of samples (again numbers by sample ID) demonstrating 7 of 8 posted/fresh sample pairs cluster closer to each other than other patient samples. Samples for one patient did not show clustering (samples 45 and 47). Red are posted samples and green are fresh samples.

### Proteomics

#### Identification of the core sputum proteome

From the entire dataset (12 participants; 24 samples), 4,088 proteins were identified based on one or more unique peptide identifications and a false discovery rate of < 1 %. To further increase confidence in protein identifications, proteins were further filtered to exclude those with > 25% missing data within a given cohort/condition (i.e. fresh or posted). This resulted in a core sputum proteome of 2,188 high confident protein assignments.

#### The sputum proteome highlights patient-specific and posted/fresh-dependent variation

Principal component analysis (PCA) highlighted clear proteome differences between fresh and posted sputum in the first two principal components without overlap, largely separating based on the PC2 axis (Fig. 6a). When visualising the PCA using patient IDs, the patients largely demonstrated comparable positioning along the PC1 axis regardless of their fresh or posted cohort categorisation (Fig. 6b). To explore the drivers of variance observed by PCA, an eigencor plot was generated by mapping patient metadata onto the first five principal components (Fig. 6c). This analysis revealed that FEV1 exhibited the strongest association with PC1 whilst sample condition (fresh vs posted) correlated most strongly with PC2, consistent with the observed group separation in the PCA (Fig. 6b). To understand more about the proteins driving these separations/associations, protein set enrichment analysis was performed (Fig. 6d). Enrichment along the PC1 axis was most strongly associated with cadherin binding and protein-folding related functions: a signature of epithelial stress and junctional remodelling while PC2 enrichment reflected differences attributable to sample condition (fresh vs posted), including vesicle-mediated transport and protein-DNA complex assembly highlighting stress-induced trafficking alterations.

**Figure 6.**
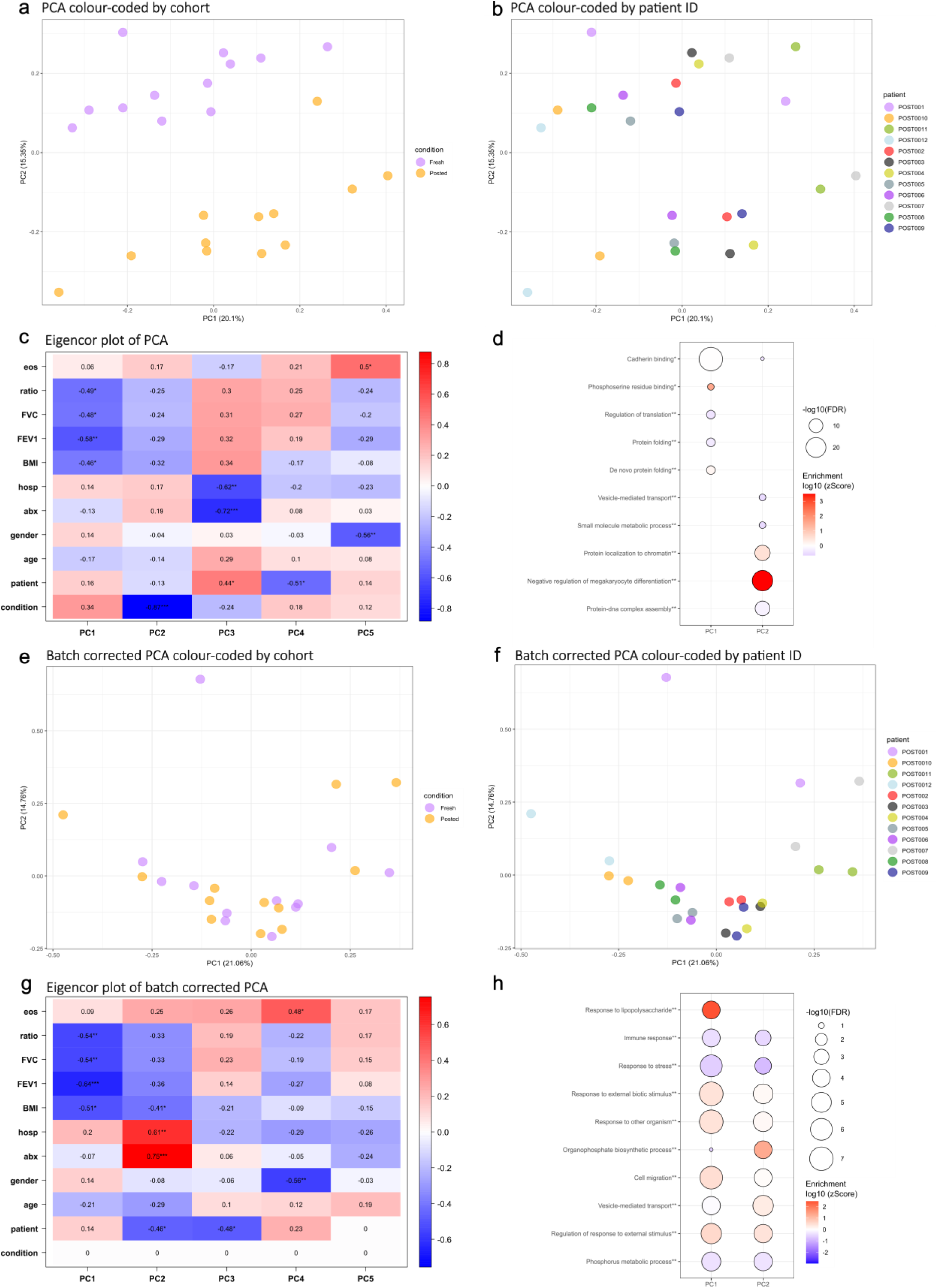
Principal component analysis of sputum protein profiles. The entire data set (12 participants; 24 samples) were subject to principal component analysis (PCA) using the core sputum proteome (2188 proteins). [A] Samples were colour-coded by cohort and [B] patient ID. [C] Correlations between the first five principal components (PC) and clinical variables were illustrated using an Eigencor plot. [D] Protein groups identified as the most enriched *molecular function and **biological process for PC1 and PC2. PCA was repeated after Limma batch-correction. [E] Samples were colour-coded by cohort and [F] patient ID. [G] Correlations between the first five PCs and clinical variables were illustrated using an Eigencor plot. [H] Protein groups identified as the most enriched *molecular function and **biological process for PC1 and PC2. **Abbreviations: eos=eosinophil count; ratio=fev1/fvc ratio; fvc=forced vital capacity; fev1=forced expiratory volume in 1 second; bmi=body mass index; hosp=hospital admissions in last 12 months; abx=courses of oral antibiotics in last 12 months; conditon=posted/fresh.**

#### Condition dependent variation is consistent and can be adjusted for using batch correction techniques

PCA was repeated following Limma adjustment, aiming to remove proteome differences attributed to sample condition i.e. postal/fresh status. Post adjustment, no clear separation was observed between fresh and posted samples, indicating fresh/posted status was driving the separation, but that the change is consistent and amenable to adjustment. (Fig. 6e). When visualising the PCA by patient ID, both samples originating from the same individual largely clustered together (Fig. 6f). Again, an eigencor plot was generated to explore the drivers of variance observed by PCA. As before, this analysis confirmed that markers of lung disease severity ( correlated most strongly with PC1, while PC2 captured variation related to exacerbation frequency (number of hospitalisations and antibiotics in the last 12M), again a key marker of disease severity. (Fig. 6g). Proteins driving the separation/associations along the PC1 axis were largely driven by immune and stress-response pathways, while PC2 enrichment was characterised by pathways liked to immune cell activation, migration and metabolic rewiring.

## Discussion

This study found postage of sputum samples allows robust assessment of *P. aeruginosa* in terms of the quantification of cultured load, amplified load, and detailed AST. Furthermore, limited effect on individual or community metrics of sputum microbiome, metabolome and proteome were observed. Collectively, these results suggest postal sputum is a valid sampling tool for a broad range of clinical and research applications.

Sputum sampling is a cornerstone of clinical management of people living with bronchiectasis and regular sampling is recommended by national and international guidelines (9, 10). Identification of pathogens, including *P. aeruginosa,* allows targeted treatment and, more broadly, sputum colour itself has been identified as an independent prognostic marker (11).

Postal sputum sampling represents a method for patient-led and potentially on demand sampling, however, while in use at some centres to date this approach has not yet been widely adopted. This is perhaps in part due to mixed results in historical studies leading to concerns regarding the ability to recover, identify, or quantify pathogens from postal sputum samples.

A key concern is the exposure of sputum to prolonged periods of ambient temperature. In the present study, we observed a mean four-day delay in sample analysis in posted samples, however, no significant impact on cultured or amplified load was discerned. In keeping with our results for *P. aeruginosa,* previous work found prolonged periods of up to six days at ambient temperatures did not impact the identification of *S. pneumoniae* (12). Similarly, no significant reduction in recovery of *H. influenzae* and *S. pneumoniae* was observed following postage (13). Furthermore, transport by post had little to no effect on the isolation of *S. pneumoniae, H. influenzae, P. aeruginosa, or Moraxella catarrhalis* from sputum from patients with chronic lung disease (14). Changes have been described in more recent studies, where samples left at ambient temperatures resulted in increased, rather than decreased, culturable load of *P. aeruginosa.* Although increases were modest in the range of 0.2 Log, this is perhaps suggestive of overgrowth at ambient temperatures (5). In contrast, a previous UK postal study of samples in CF found no impact of postage on the bacterial abundance of five important CF pathogens, including *P. aeruginosa* (4), supporting our results here suggesting that temperate conditions of postage do not have meanginful impact on culturable bacterial load.

Our study expands on these previous works in a number of key areas. First, we explored impact of postage on AST in addition to recovery and quantification of *P aeruginosa,* the major pathogen in bronchiectasis. In clinical utility, the recovery rather than quantification of *P. aeruginosa* holds more relevance in that different loads do not currently alter management significantly. However, overgrowth does have the potential to impact clinical decision making particularly if it resulted in distorted AST. We therefore characterised the extended AST of ten isolates per positive sample (140 isolates in total) to 12 antibiotics, including newer infrequently used anti-Pseudomonals. To the best of our knowledge, ours is the first study to explore this and we found little evidence of changes in AST profile with excellent agreement across samples, supporting our cultured load results and suggesting AST profiles derived from postal sputum samples are valid for a comprehensive range of clinically relevant antibiotics.

Next, we evaluated the utility of postal sampling and the impact of DNA/RNA shield for a range of research applications including oral microbiome, metabolome, and proteome. Our 16S microbiome analyses identified a Pseudomonas taxon as the most abundant, an expected finding given the inclusion criteria of the study. We were able to detect marked differences in Pseudomonas abundance between those samples that were culture-negative and culture positive; however we found no consistent effect of postage on the abundance of Pseudomonas. Similarly, we found no changes in community composition in terms of diversity metrics or individual taxa suggesting postal samples are valid for microbiome analyses in this setting.

To the best of our knowledge, this study is the first to explore the effects of postage on metabolites present in sputum. Similar results were observed across metabolome analyses, with distinct metabolomes identified between patients but little apparent effect of postage. The use of DNA shield did negatively impact on the metabolomic profiles acquired thus suggestive sputum samples for metabolomic analyses should be collected and stored without the use of DNA shield.

Conversely, we did identify differences in the sputum proteome in posted samples, again to our knowledge the first time this has been explored. In total, 408 proteins were significantly differentiated between posted/fresh samples (resampled P <0.05) and differential proteins were largely related to intracellular metabolism pathways (Supp. Figure 3). Postage/Fresh status accounted for the vast majority of PCA variation in PC2 axis, which accounted for 15.3% of all variation. With disease severity characteristics, e.g. lung function and BMI, driving most of the variation in PC1 (20.1%).

Taken together the lack of change in the metabolome but change in proteome may be consistent with extracellular leakage from cell death in the postal samples, rather than any up/downregulation of metabolic pathways. Importantly, the effect of postage was consistent and bioinformatic Limma batch-correction was able to effectively adjust for postage related proteomic change. This holds particular relevance to future clinical research and study design, by suggesting that an initial paired posted/fresh sample could be followed by longitudinal postal samples. The strategy could have broad applications in clinical trials but also in helping understand dynamic transitions from stable to exacerbation state in chronic lung infection.

A key methodological consideration for culture-independent analyses is freeze-thaw effects and the utilisation of a preservative, such as DNA/RNA shield. A singular freeze-thaw cycle at -80°C resulted in a small but consistent increase in the amplified load of *P. aeruginosa*, which is perhaps unsurprising given the ability of temperature extremes to lyse bacterial cells. In the present study, the effects of multiple freeze-thaw cycles were not determined, however, previous studies indicate that sputum microbial communities are robust for up to six freeze-thaw cycles (15). The use of DNA/RNA shield had no significant impact on the amplified yield of *P. aeruginosa* in fresh or posted sputum and similarly, had no effect on alpha or beta diversity metrics in the 16S microbiome. However, DNA/RNA shield did significantly hinder metabolomic and proteomics analysis rendering the results uninterpretable in those samples.

### Limitations

As the present study recruited patients from a single clinic and laboratory work was conducted at a single institution, it is recognised that a multi-centre approach would allow a more accurate representative population, at both microbiological and disease level. Furthermore, the present study focused solely on patients thought to be chronically infected with *P. aeruginosa* and as such, the effectiveness of postal samples for the isolation of other pathogens remains to be explored. However, previous postal studies investigating *H. influenzae* (*13*)*, S. pneumoniae* (12), and *M. catarrhalis* (14) found little to no effect of postage on bacterial isolation. Furthermore, a retrospective study of over 34,000 sputum samples demonstrated the isolation of clinically important fungal species following postage at ambient temperature, concluding that samples should not be rejected due to a timelapse between sample collection and culture (12). This suggests that postal sputum samples may be suitable for the isolation of other clinically important pathogens, however, further work is needed to confirm applicability. The present study took place during winter (December-February), where average outside UK temperatures were approximately 5°C. It is unknown how seasonal temperatures during transit could have impacted bacterial abundance; *P. aeruginosa* can proliferate at room temperature, and significant increases in bacterial density have been described in sputum stored at room temperature (5). Finally, it is acknowledged that small sample size may limit our power to detect certain biological changes, particularly at an omics level, with lowly abundant taxa, proteins, or metabolites potentially being underrepresented.

## Conclusion

In summary, this study compared the impact of fresh and posted sputum sampling on the recovery and quantification of *P. aeruginosa*, antibiotic susceptibility as well as more broadly for multi-omic analyses. Postal samples appear valid for a range of clinical and research applications in this population. Future work determining their utility in early diagnosis and eradication protocols, and longitudinal decentralised research, incorporating different climates and travel distances is warranted.

## Data Availability

All data produced in the present study are available upon reasonable request to the authors.

## Supplementary Materials

### Supplement Contents

1. Supplementary Figures
2. Supplementary methodologies

### 1. Supplementary Figures

**Supp. Figure 1.**
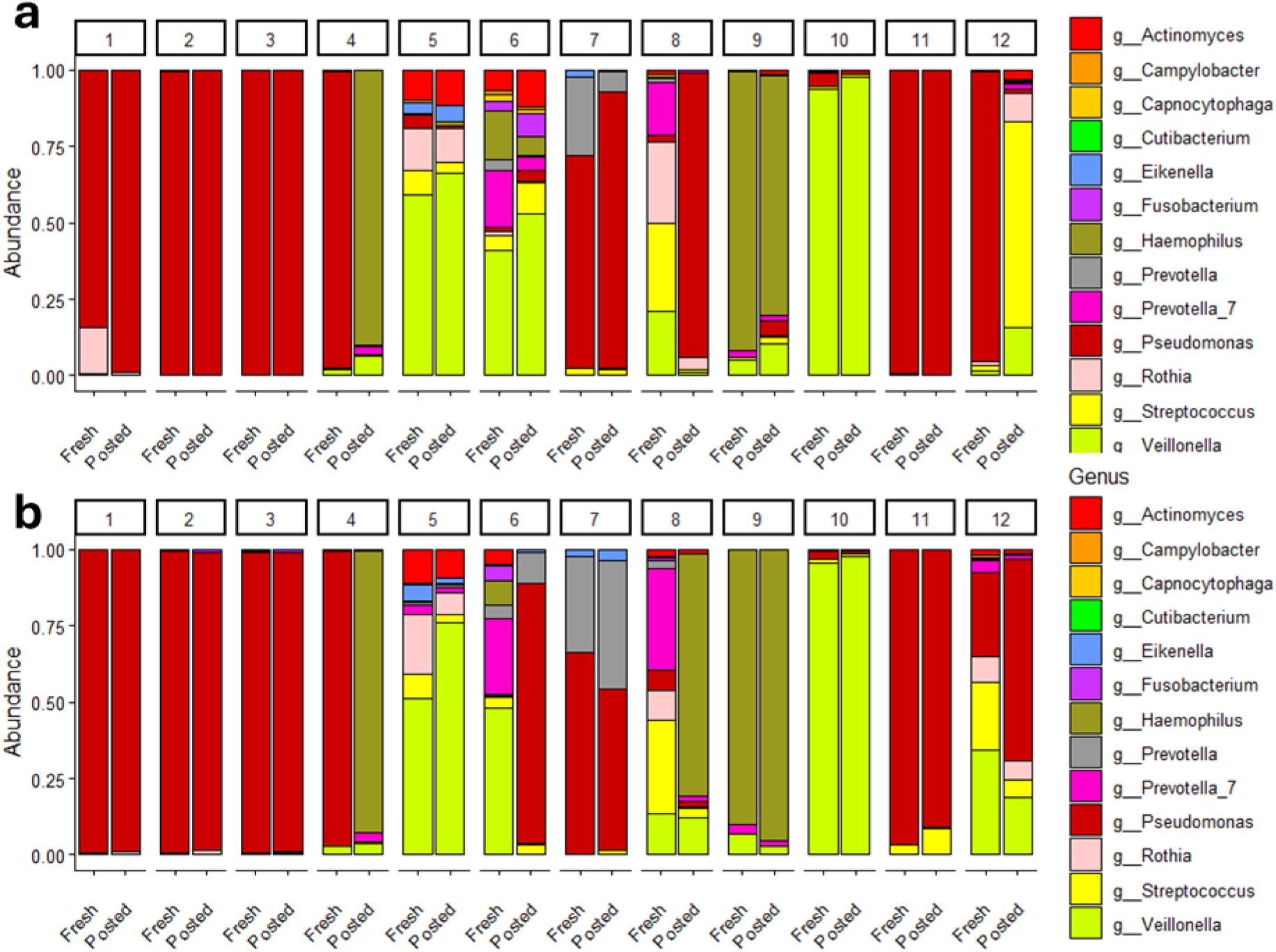
Abundance plots for top genus identified in fresh and posted samples, without [a] and with [b] the addition of DNA shield. No taxa were differentially abundant between fresh and posted samples.

**Supp. Figure 2.**
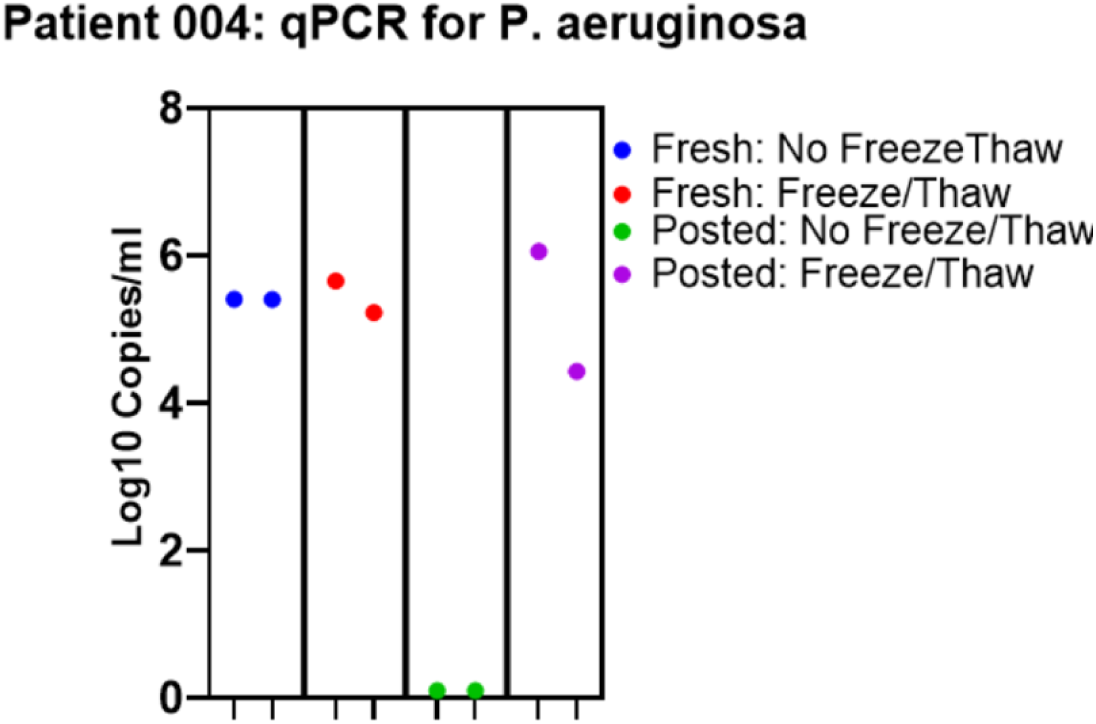
Quantification by qPCR of copies of *P. aeruginosa* in patient with discrepancy in abundance of *Pseudomonas* reads between fresh and posted samples. No *P. aeruginosa* copies were detected in the posted samples when analysed prior to freezing, however after a freeze-thaw cycle copies were detected in similar range to that in the same-day fresh samples.

**Supp. Figure 3.**
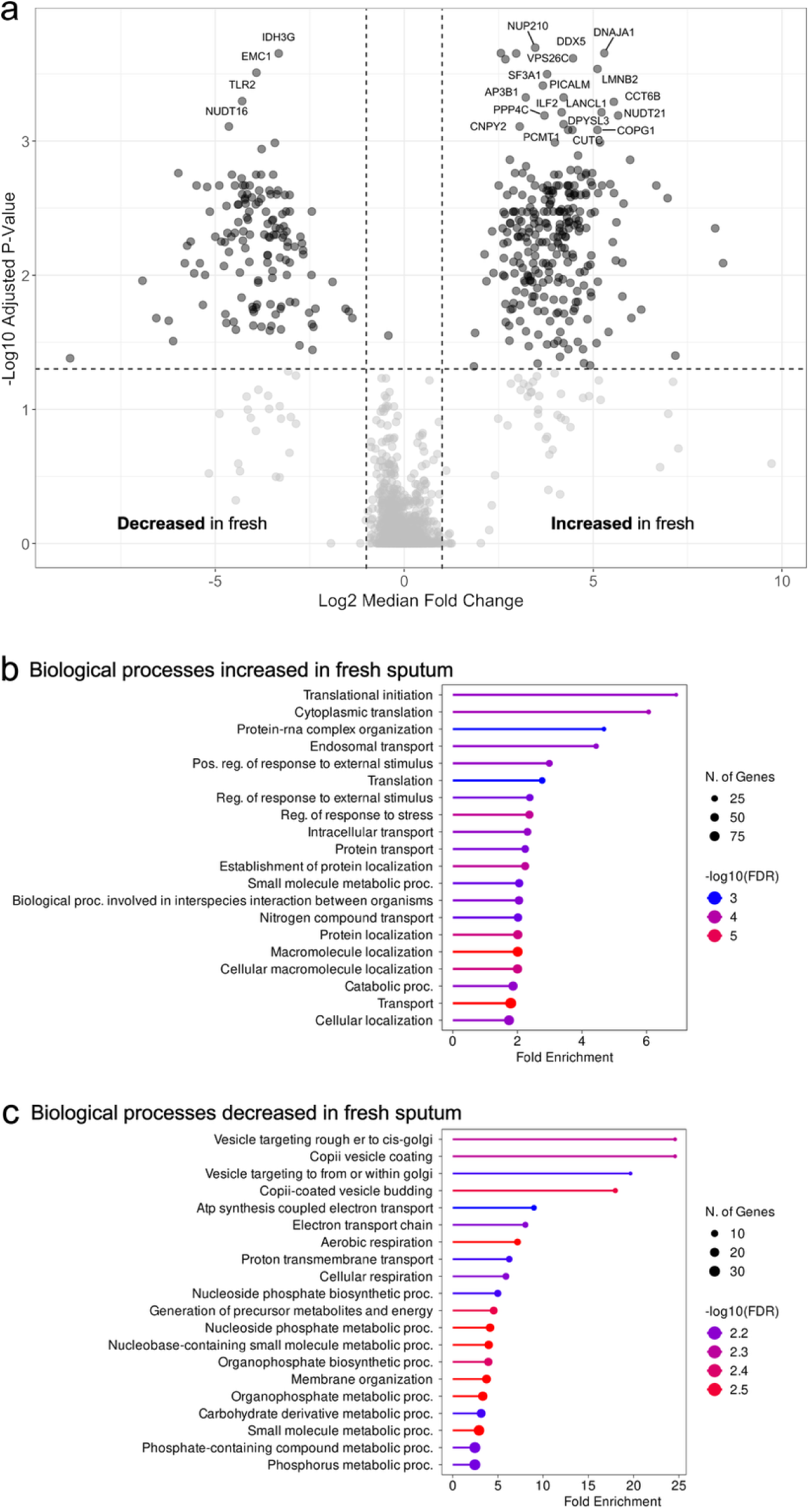
[a] Up- and downregulated proteins, expressed in fresh and posted sputum. Biological processes increased [b] and decreased [c] in fresh sputum, compared to postal sputum. In total, 408 proteins were significantly differentiated between posted/fresh samples (resampled P <0.05) and differential proteins were largely related to intracellular metabolism pathways.

### 2. Supplementary Methods

#### Sample Storage

Culture and initial DNA extraction was performed on fresh unfrozen processed sputum samples. Following this, processed sputum samples were stored at -80°C and extracted DNA was stored at - 20°C. Ten bacterial isolates from each positive culture plate were stored in 96-well flat bottomed plates at -80°C, by adding a singular colony to 200 µL glycerol stock (5% glycerol LB broth) in each well. Isolates were recovered for susceptibility testing by streaking 5 µL of glycerol stock onto a Pseudomonas-selective agar plate.

#### 16S Microbiome Sequencing

Primers described by Caporaso et al (2011 PNAS 108, 4516-4522) were used to amplify the V4 region of 16S. 2µl of submitted sample entered a first round PCR with conditions 20sec at 95°C, 15sec at 65°C, 30sec at 70°C for 20 cycles then a 5min extension at 72°C. KAPA HiFi HotStart ReadyMix used for PCR reaction, primer design below:

F: 5’ACACTCTTTCCCTACACGACGCTCTTCCGATCTNNNNNGTGCCAGCMGCCGCGGTAA3’

R: 5’GTGACTGGAGTTCAGACGTGTGCTCTTCCGATCTGGACTACHVGGGTWTCTAAT3’

The Primer design incorporates a recognition sequence to allow a secondary nested PCR process. Samples are first purified with SPRI Beads before entering the second PCR performed to incorporate Illumina sequencing adapter sequences containing indexes (i5 and i7) for sample identification. 15 cycles of PCR were performed using the same conditions as above for a total of 35 cycles. Samples were purified using SPRI Beads before being quantified using Qubit and assessed using the Fragment Analyzer. Successfully generated amplicon libraries were taken forward.

Final libraries were pooled in equimolar amounts using the Qubit and Bioanalyzer data. The quantity and quality of each pool was assessed by Bioanalyzer and subsequently by qPCR using the Illumina Library Quantification Kit from Kapa (KK4854) on a Roche Light Cycler LC480II according to manufacturer’s instructions. Briefly, a 10 µl PCR reaction (performed in triplicate for each pooled library) was prepared on ice with 6 µl SYBR Green I Master Mix and 2 µl diluted pooled DNA (1:1000 to 1:100,000 depending on the initial concentration determined by the Qubit® dsDNA HS Assay Kit). PCR thermal cycling conditions consisted of initial denaturation at 95°C for 5 minutes, 35 cycles of 95°C for 30 seconds (denaturation) and 60°C for 45 seconds (annealing and extension), melt curve analysis to 95°C (continuous) and cooling at 37°C (LightCycler® LC48011, Roche Diagnostics Ltd, Burgess Hill, UK).

Following calculation of the molarity using qPCR data, template DNA was diluted to 10.5pM and denatured for 5 minutes at room temperature using freshly diluted 0.1 N sodium hydroxide (NaOH) and the reaction was subsequently terminated by the addition of HT1 buffer. To improve sequencing quality control 15% PhiX was spiked-in. The libraries were sequenced on the Illumina® MiSeq platform (Illumina®, San Diego, USA), using V2 chemistry, generating 2 x 250 bp paired-end reads.

#### Metabolomics

##### Metabolite extraction

Sputum samples (150 µL) were pipetted into 2 mL Eppendorf tubes. To the aliquots, 450 µL of methanol was also added for the purpose of protein precipitation and extraction of metabolites. The mixture was then vortexed (15 seconds) and centrifuged (17,000 *xg*, 15 minutes, 4°C). After centrifugation, 450 μL of the supernatant was aliquoted into a different 2 mL Eppendorf tube. For UHPLC-MS analysis all samples were analysed directly. For GC-MS analysis. all samples underwent vacuum lyophilization overnight at 35°C. A blank sample was prepared using the method described above but in the absence of a biological sample. Pooled QC samples were prepared by pooling 200 µL aliquots of all biological samples and 150 µL aliquots were prepared as defined above for GC-MS and UHPLC-MS analysis.

##### Gas chromatography-mass spectrometry analysis

For derivatization of dried sputum samples, 200 μL of a 20mg.mL^-1^ methoxyamine in pyridine solution was prepared. 200 µL of the methoxyamine in pyridine solution was added. The samples were then subjected to incubation at 80°C for 20 minutes. 100 µL of N-Methyl-N-(trimethylsilyl)trifluoroacetamide (MSTFA) was added to each tube and incubated at 80°C for a further 20 minutes. After incubation, the samples were allowed to cool to room temperature, were centrifuged (17,000 *xg*, 15 minutes, 4°C) and then aliquots were transferred to a GC vial.

All samples were analysed using an Agilent 8890 Gas Chromatograph (Agilent Technologies, Stockport, UK) coupled with a LECO Pegasus BT TOF mass spectrometer (Leco, Stockport, UK). QC samples were analysed for the first five injections and then every sixth injection. The last two injections were also QC samples. 1 μL split injections (1:10 split) were made at 280 °C with a Gerstel LPAL3 DRV autosampler, onto a 30m × 0.25mm × 0.25 μm RXi-5MS column (Restek) through an Agilent Split/Splitless injector equipped with a 4 mm split liner. Helium carrier gas was set at a 1.4 mL.min^-1^ constant flow; oven temperature programme was held at 40 °C for 0.7 min, then a linear temperature ramp from 40 to 300 °C at 20 °C.min^-1^, with a hold temperature of 300 °C for 3 min. The column was led directly into the ion source of the mass spectrometer held at 250 °C via a transfer line held at 280 °C. Data were acquired at 20 Hz, over the m/z range 45–500Da, with a 275 s solvent delay prior to acquisition. A retention index mixture (C_8_-C_30_ n-alkanes) was analysed as the first injection to define retention indices across the run.

Raw instrument data were pre-processed applying the ChromaTof software (LECO; v5.55) using a minimum signal-to-noise of 100 and a stick count > 10. A QC sample and a biological sample were processed and a reference file constructed. The reference file was applied to process all sample files. The peak area was reported for the Quan ion. These data representing normalised peak lists were exported in.txt format. Most metabolites were putatively annotated (MSI level 2 or 3) according to The Metabolomics Standards Initiative reporting standards (Sumner et al., 2007) by comparison of mass spectra acquired from the samples with those present in a mass spectral library (NIST23). Processed data were manually integrated in Microsoft Excel® (.xls) worksheets for further data analysis.

##### Ultra High Performance Liquid chromatography-mass spectrometry analysis

The samples (maintained at 4°C) were analysed applying a Vanquish binary pump H system coupled with a heated electrospray Orbitrap Exploris 240 mass spectrometer (Thermo Fisher Scientific, MA, USA). Sample extracts were analysed using a Hypersil GOLD C_18_ column (100 x 2.1mm, 1.9 μm; Thermo Fisher Scientific, MA, USA). For positive and negative ion modes mobile phase A was 60% acetonitrile/40% water (10 mM ammonium formate, 0.1% formic acid) and mobile phase B was 85.5% propan-2-ol /9.5% acetonitrile/5% water (10 mM ammonium formate, 0.1% formic acid). The gradient elution applied was: t=0.0, 20% B; t=1.5, 20% B; t=3.0, 25% B; t=9.2, 100% B; t=10.5, 100% B; t=12.5, 20% B; t=15.0, 20% B. All changes were linear (curve = 5) and the flow rate was 0.40 mL.min^-1^. Column temperature was 55°C and injection volume was 2 μL. Data were acquired in positive and negative ionisation modes separately in the m/z range of 150 –2000 with a mass resolution of 120,000 (FWHM at *m/z* 200). Ion source parameters applied were sheath gas = 40 arbitrary units, aux gas = 8 arbitrary units, sweep gas = 1 arbitrary units, spray voltage = 3.2kV (positive ion mode) and 2.7kV (negative ion mode), vaporizer temperature = 320°C and ion transfer tube temperature = 250 °C. All samples were collected as MS1 data in the profile mode applying: scan time = 100ms, microscans = 1, RF lens = 60% and normalised AGC target=100%. For peak annotation purposes, MS/MS data were collected in the “Data dependent mode” setting on five QC samples analysed as injections 7-11 in each batch over different *m/z* ranges (150-500 *m/z*; 500-710 *m/z*; 700-860 *m/z*; 850-1010 *m/z* and 1000-2000 *m/z*) using stepped normalized collision energies of 20/50/130% (negative ion mode) and 20/40/100% (positive ion mode). MS/MS data were applied with the number of dependent scans = 3, a mass resolution of 15,000 (FWHM at *m/z* 200) and an isolation width = 3 *m/z*. Orbitrap Exploris 240 Tune application software controlled the instrument.

Data files (in the .RAW file format) were converted to .mzML file formats applying msConvert in Proteowizard [1]. Data (mzML format) for each sample were processed applying the R package XCMS to construct a single data matrix for all samples (metabolite features as rows and samples as columns) for quality assessment and statistical analysis [2]. XCMS applied three steps. Step1: Peak detection (“findChromPeaks”) using the “centWave” algorithm was employed with parameter settings of m/z deviation = 25 ppm, peakwidth = 5 ∼ 20, snthresh = 10, prefilter = 3 ∼ 100, mzCenterFun = "wMean") and mzdiff = 0.001. Step 2: Alignment (“adjustRtime”) was applied to perform retention time correction (alignment) between chromatograms of different samples using the Obiwarp method with parameters of binSize = 1, gapInit = 0.4) and gapExtend = 2.4. Step 3: Peak grouping (“groupChromPeaks”) was performed to group the chromatographic peaks within and between samples. The sample/replicate category group, such as “sample”, “QC” and “blank” was used as peak group information.

##### Data quality assessment

The quality of data was calculated so to remove poor quality data. For QC sample data, metabolite peaks were removed if they were detected in < 70% of the QC samples or the relative standard deviation (RSD) > 30%.

##### Univariate and multivariate analysis

MetaboAnalyst v6.0 [3] a web-based platform was applied for metabolomics data analysis. The processed data sets from GC-MS and LC-MS, which included peak intensity measurements for each detected metabolite, were uploaded to the platform in .csv format.

For univariate analysis - missing values were not imputed, data were sum normalised and log2 transformed. Paired t-test analysis was performed of samples with the paired samples being fresh and posted samples for the same subject. Correction for multiple testing was applied using the Benjamini-Hochberg method. Hierarchical Clustering Analysis was performed with Euclidean distance measure and the Ward clustering algorithm.

For Principal Component Analysis - missing values were imputed (kNN), data were sum normalised, log2 transformed and Pareto scaled.

#### Proteomics

##### Proteomic sample preparation

Processed sputum samples, processed as previously described, were thawed from storage at -80°C. SP3 tryptic digestion: Sputum samples were diluted in 25 mM ammonium bicarbonate (AmBic) to a final volume of 75 μL and normalised to a total protein concentration of 25 µg. Protein denaturation was performed by the addition of 10 % (w/v) SDS to a final concentration of 1 % (w/v) before incubating at 95 °C for 10 minutes. Cysteine reduction was performed by addition of DTT to a final concentration of 4 mM, before incubating at 60 °C for 10 minutes. Iodoacetamide (IA) was added to a final concentration of 14 mM to alkylate the free sulfhydryl groups on the cysteine residues. After incubation for 30 minutes at room temperature in the dark additional DTT was added to a vinal concentration of 7 mM to quench excess IA. Next, 10 µL of a 50 ug/µL solution of washed and combined Sera-Mag SP3 beads (E3 and E7) were added followed by 182.8 µL of 100% (v/v) ethanol to bind the protein to the beads. The beads were gently mixed by pipetting and incubated for 30 minutes at room temperature before a magnetic rack was used to pellet the beads, and the supernatant was removed. Samples were washed six times with 400 µL of 80% (v/v) ethanol, leaving the sample to stand for 5 minutes before removing the wash solution. After the final wash, SP3 beads were dried for 10 minutes in a speedVac (Centrifuge: UNIVAPO – 150 ECH, Cooling unit: UNICRYO MC2L -60 °C, Vacuum pump: UNIVAC DQ4) to remove trace ethanol. Protein digestion was performed by the addition of 100 µL of 0.01 µg/µL trypsin and incubating at 37 °C overnight (∼16 hours). Digestion was stopped by the addition of trifluoroacetic acid (TFA) to a final concentration of 0.5% (v/v) and incubated for 45 minutes at 37 °C. The samples were centrifuged to remove particulates for 15 minutes at 13,000 x g. The samples were then subject to LC-MS/MS analysis.

##### Proteomic based LC-MS/MS analysis

Peptides were analysed using an Evosep One (Evosep Biosystems, Denmark) coupled online to a hybrid trapped ion mobility spectrometry - quadrupole time of flight mass spectrometer (timsTOF HT, Bruker Daltonics, Bremen, Germany) with a modified nano-electrospray ion source (CaptiveSpray, Bruker Daltonics). EvoTips (EvoSep, Odense, Denmark) were prepared as per the manufacturer’s instructions. Briefly, EvoTips were washed with 20 μL 0.1% (v/v) FA (ACN) and centrifuged at 800 g for 1 minute. The tips were then conditioned using 100% (v/v) isopropyl alcohol before washing with 20 μL 0.1% FA (v/v) (H2O) and centrifugation at 800 g for 1 minute. Next, 2 μL of sample was added to the tip and the centrifugation step was repeated. The tips were then washed once more with 0.1% FA (v/v) (H2O), and centrifuged. To prevent the tips from drying out before analysis, 200 μL 0.1% FA (v/v) (H2O) was then added. Samples were analysed using the following method https://www.evosep.com/wp-content/uploads/2021/05/AN-009A-30SPD.pdf. The mass spectrometer was operated in data independent PASEF mode. The dia-PASEF method was defined with a m/z range of 400-1201 and a mobility range of 1/ K0 = 0.6-1.6 Vs/cm2 using equal ion accumulation and ramp times of 100 ms in the dual TIMS analyzer. The collision energy was lowered as a function of increasing ion mobility from 59 eV at 1/K0 = 1.4 Vs/cm2 to 20 eV at 1/K0 = 0.6 Vs/cm2. The dia-PASEF window scheme consisted of 32 isolation windows of 26 m/z width (cycle time: 1.8 s) with 1 Da mass overlap and 1 Ion Mobility Window.

##### Proteomic data analysis

Analysis of raw LC-MS/MS data was performed using Spectronaut V19.2. Data were searched against the Uniprot human database and filtered to contain only one protein per gene. Data were searched using trypsin as the specified protease with a dynamic modification of oxidation (M) and acetyl (protein N-term), and with the static modification of carbamidomethylation (C), limited to 1 missed cleavage. Peptide mass and fragment mass tolerances were defined at ±25 ppm and ±0.05 Da, respectively.

## Notes

### Competing Interest Statement

The authors have declared no competing interest.

### Author Declarations

Ethics committee of the Liverpool University Biobank (Health Research Authority (18/NW/0771)) gave ethical approval for this work.

### Funding

- Asthma and Lung UK: DPP23\20
- Cystic Fibrosis Trust

